# Hydrating on the synthetic rainbow: Prevalence of synthetic food dyes in hydration drinks

**DOI:** 10.64898/2026.03.11.26348123

**Authors:** Angel Castro Polvorosa, Lilly Bacock, Thomas Shumway, Staci Denham, Jolene Coverston, Rebecca Bevans

**Author notes:** Correspondence concerning this article should be addressed to Angel Castro Polvorosa (AP). We have no conflict of interest to disclose.

## Abstract

Synthetic food dyes (SFDs) have been associated with neurobehavioral symptoms in children, including hyperactivity, inattention, anxiety, and irritability. Although often associated with candy and snack foods, SFDs are also present in hydration beverages commonly consumed by children during illness and physical exertion. This study examined the prevalence of SFDs in hydration drinks by target age group and product type. Ingredient information was collected for 102 hydration beverages from a single retail in Carson City, Nevada in 2024. Products were categorized as pediatric-marketed, adult-marketed, or marketed to all ages. Of 24 pediatric-marketed drinks, 21 (87.5%) contained at least one synthetic dye, most commonly Red 40 (66.7%), Blue 1 (61.9%), and Yellow 6 (19.0%). Among six adult-marketed drinks, two (33.5%) contained dyes, primarily Blue 1 and Red 40. Of 72 all-age products, 20 (27.7%) contained synthetic dyes. Overall, 43 of 102 drinks (42.2%) contained at least one SFD, with Blue 1 (51.2%), Red 40 (44.2%), and Yellow 6 (23.3%) being most prevalent. Hydration beverages are often perceived as health-supportive and are frequently consumed during physiological stress. Given prior evidence linking SFDs to behavioral effects, the high prevalence of dyes in pediatric hydration products warrants clinical awareness.

## Introduction

Synthetic food dyes (SFDs) have been known to be a predominant choice in the food industry for several decades. These dyes are primarily derived from petroleum and coal tar [1]. Azo dyes, such as Tartrazine (FD&C Yellow 5), Sunset Yellow (Yellow 6), Erythrosine (Red 3), and Allura Red (Red 40), along with triarylmethane dyes, including Brilliant Blue (Blue 1) and Fast Green (Green 3), are widely utilized to enhance the visual appeal of food products. While these synthetic dyes effectively produce vibrant colors, they are classified as xenobiotics, substances foreign to the human body, and have been implicated in adverse health effects, including chronic inflammation, microbiome disruption (dysbiosis), DNA damage, and endocrine disruption [1–15].

A substantial body of research, including both animal studies and double-blind clinical trials, has investigated the potential effects of synthetic food dyes on physiological and behavioral outcomes, notably in children. From these studies, they have established a correlation between synthetic dye consumption and neurobehavioral disturbances, including hyperactivity and aggression [16–25]. However, there remains a significant gap in the literature regarding SFDs. Notably, only a single published study has examined the prevalence of synthetic dyes in foods specifically marketed toward children [18]. However, no studies to date have investigated the presence of synthetic dyes in hydration beverages intended for infants, children, and adults. Addressing this research gap, the present study aims to systematically assess the prevalence of synthetic dyes in hydration drinks, thereby contributing to a more comprehensive understanding of exposure risks across different age groups.

## Methods

Data collection consisted of photographs of hydration drinks in liquid, powder, caffeinated, and pediatric drink formulations for both adults and children. A total of 102 hydration drinks were collected from one Consumer Value Store (CVS) in Carson City, Nevada (89706) in 2024. All 102 hydration drinks were photographed front and back, then collaged into one singular photo using a photo application, InShot. Documentation of the presence of synthetic food dyes (SFD), formulation type, presence of added sugar, and perceived health status was entered into a Google spreadsheet. Further analysis of SFDs was conducted to analyze the specific type of dye and how many the product contained. Products marketed towards infants and children were identified based on labeling terminology like “Pedialyte” or “Pediatric.” Hydration products containing caffeine were documented as adult-only hydration products. “Healthier” hydration products were also identified based on marketing terminology that promoted electrolyte-balanced, immune-booster, or faster hydration, such as “more hydrating”, “33% more electrolytes”, and “advanced care with immune support”.

## Results

From our analysis of 102 hydration products, a total of 43 (42.2%) products contained at least one synthetic food dye (SFD), while 14 (32.5%) products contained two SFDs, and 59 (58.5%) contained no SFDs. Of the 43 products containing SFDs, 32 (74.4%) were in liquid form, while 11 were in powder form. Out of the 59 products that were free of synthetic dyes, 21 (35.6%) were liquids, with 38 (64.4%) being in powder form.

**Figure 1.**
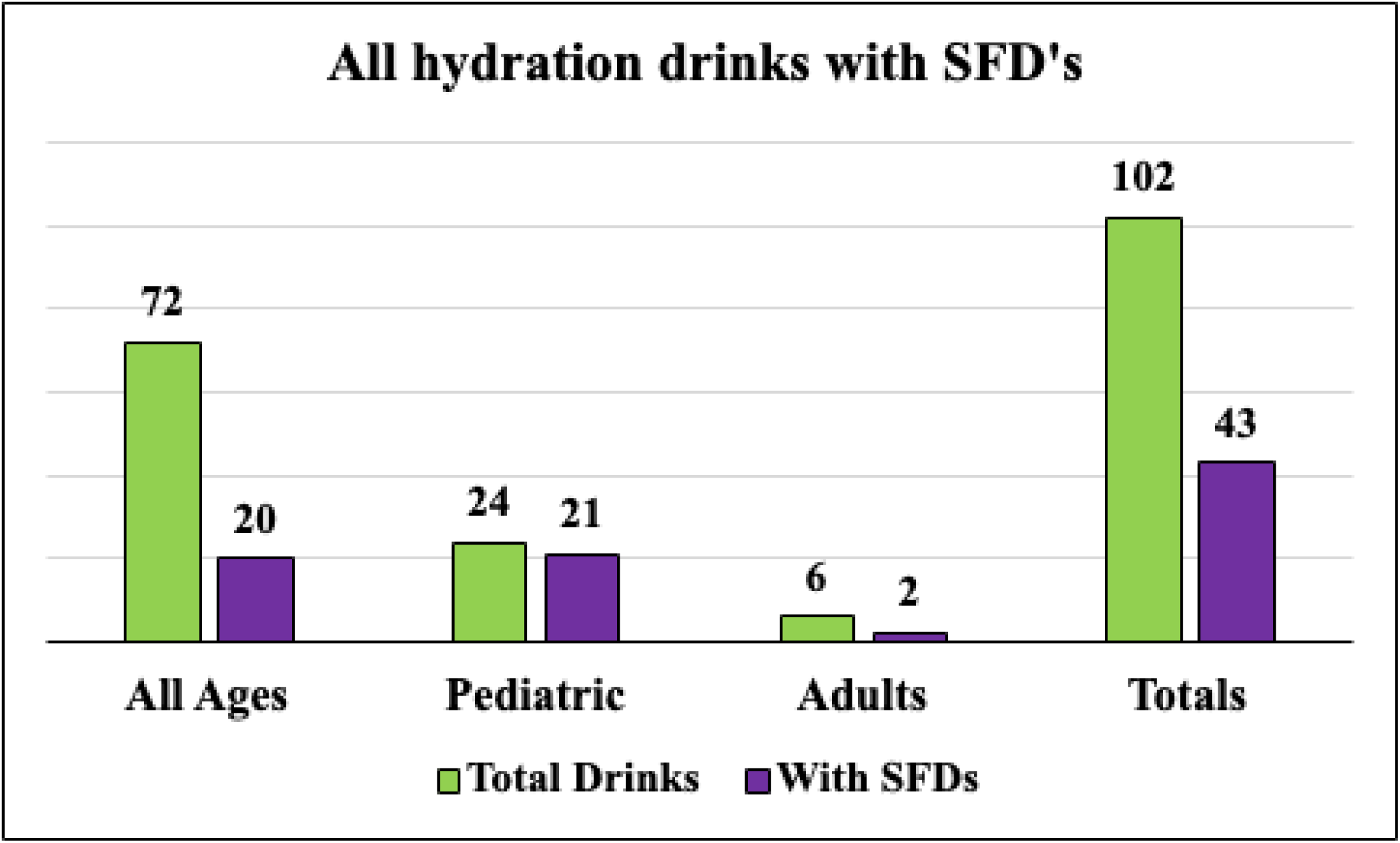
Breaks down the 102 hydration drinks into three age-related marketing categories with their own sub totals in comparing how many of those products contain SFDs.

An overall review of colors used demonstrated that Red 40, Blue 1, and Yellow 6 were the most common synthetic dyes identified among all 102 hydration drinks. These findings are consistent with prior research [18]; however, Red 40 was notably more prevalent in beverages marketed to infants and children than in those marketed to adults. In adult drinks, it showed Blue 1 to be the most common with Red 40 being second. The disproportionate presence of synthetic dyes in child-targeted products (87.5%) compared to all-age hydration products (38.4%) suggests a higher likelihood of exposure in pediatric populations. It is concerning, given that infants and children have lower body masses and developing nervous systems, both of which may heighten their sensitivity to xenobiotic substances such as synthetic food dyes.

**Figure 2.**
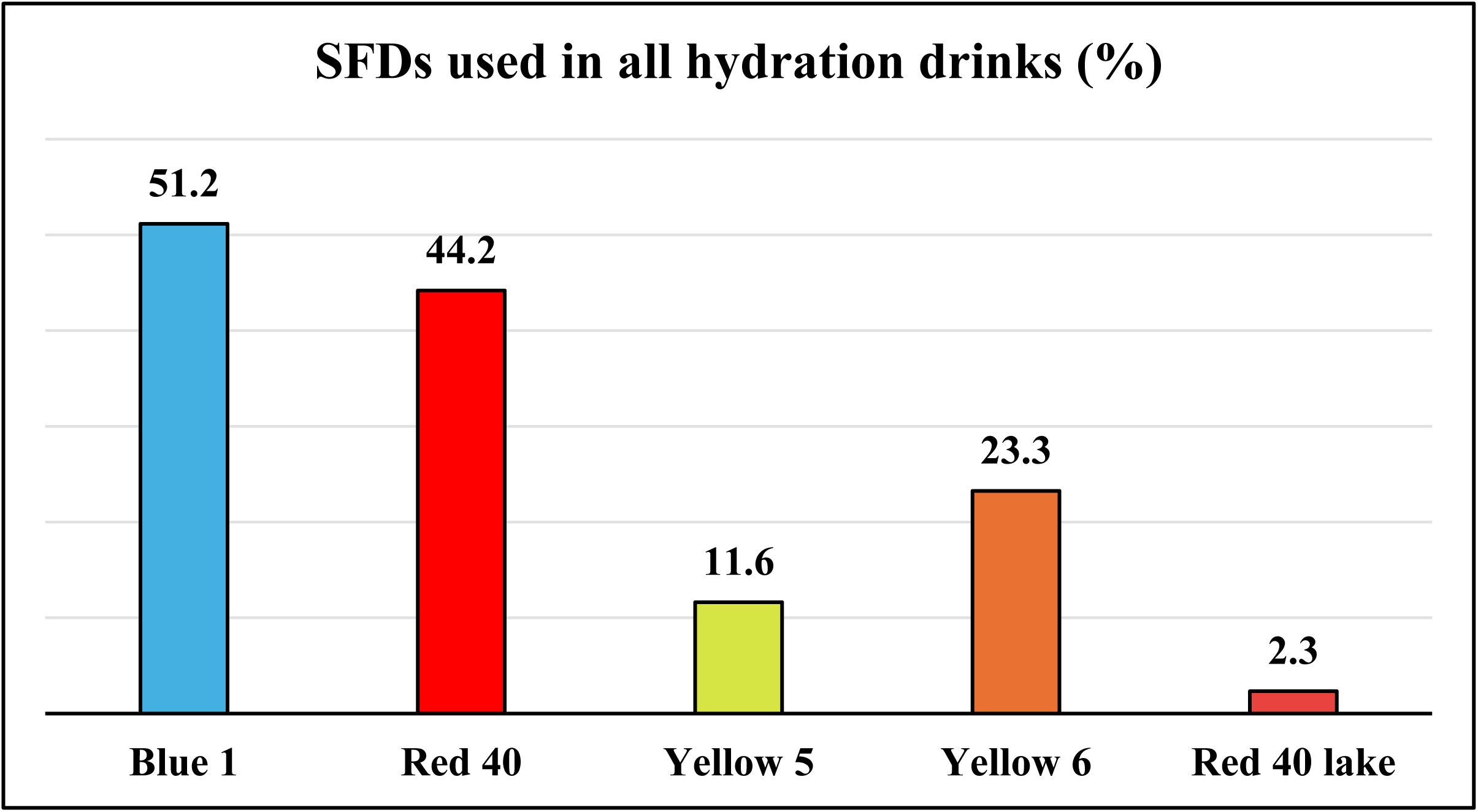
Illustrates all dyes commonly found in hydration products with synthetic dyes at CVS in Carson City, NV.

**Figure 3.**
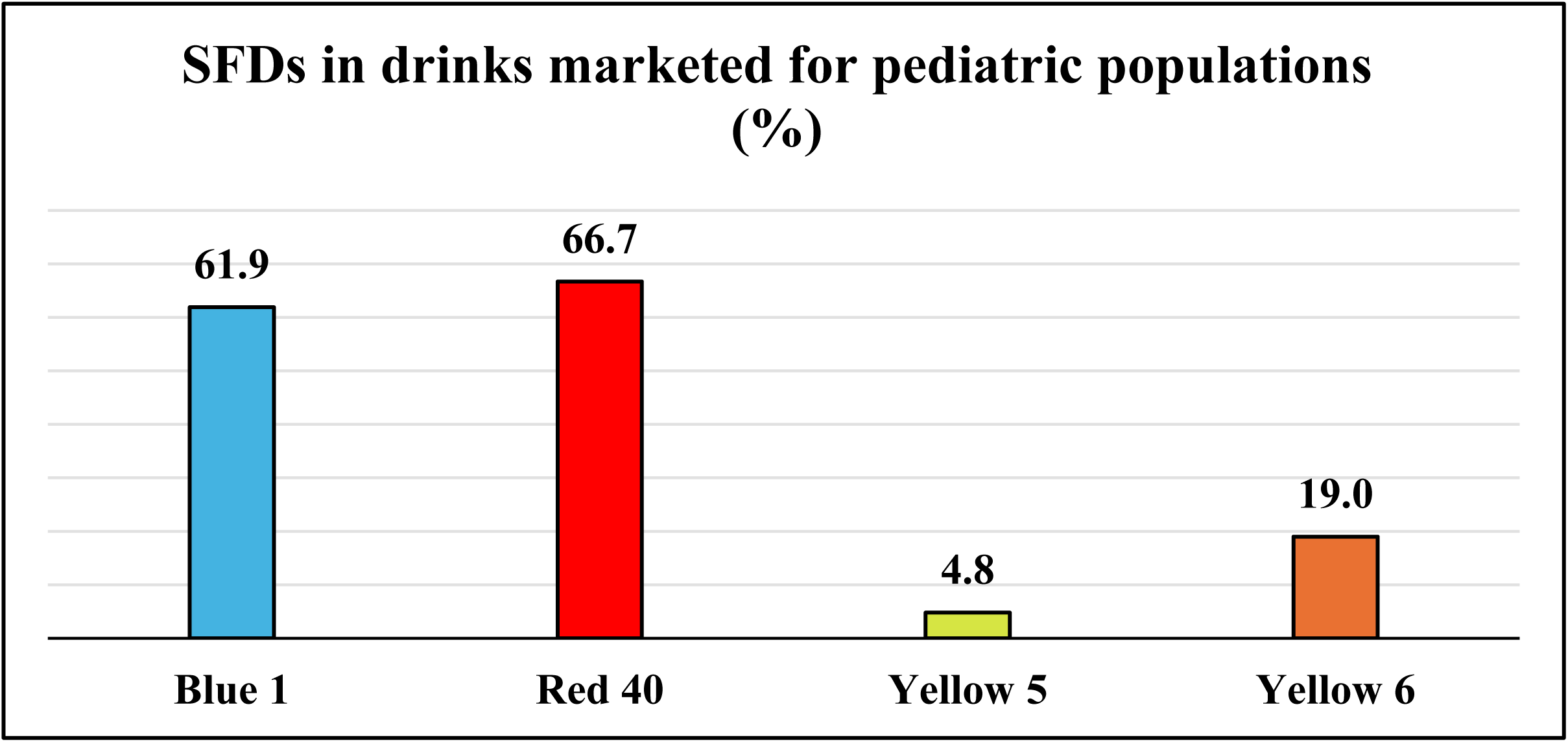
Illustrates the most frequent dyes in hydration products for pediatric population at CVS in Carson City, NV.

The study observed an association between the presence of added sugars and synthetic food dyes in hydration beverages. Of the 102 products, 82 (81.3%) contained added sugar; among those, 35 (43.2%) also contained SFDs. In contrast, of the 19 products without added sugar, only 8 (19.3%) contained SFDs. This pattern suggests that beverages formulated primarily for taste appeal, rather than solely for hydration, are more likely to contain artificial additives.

**Figure 4.**
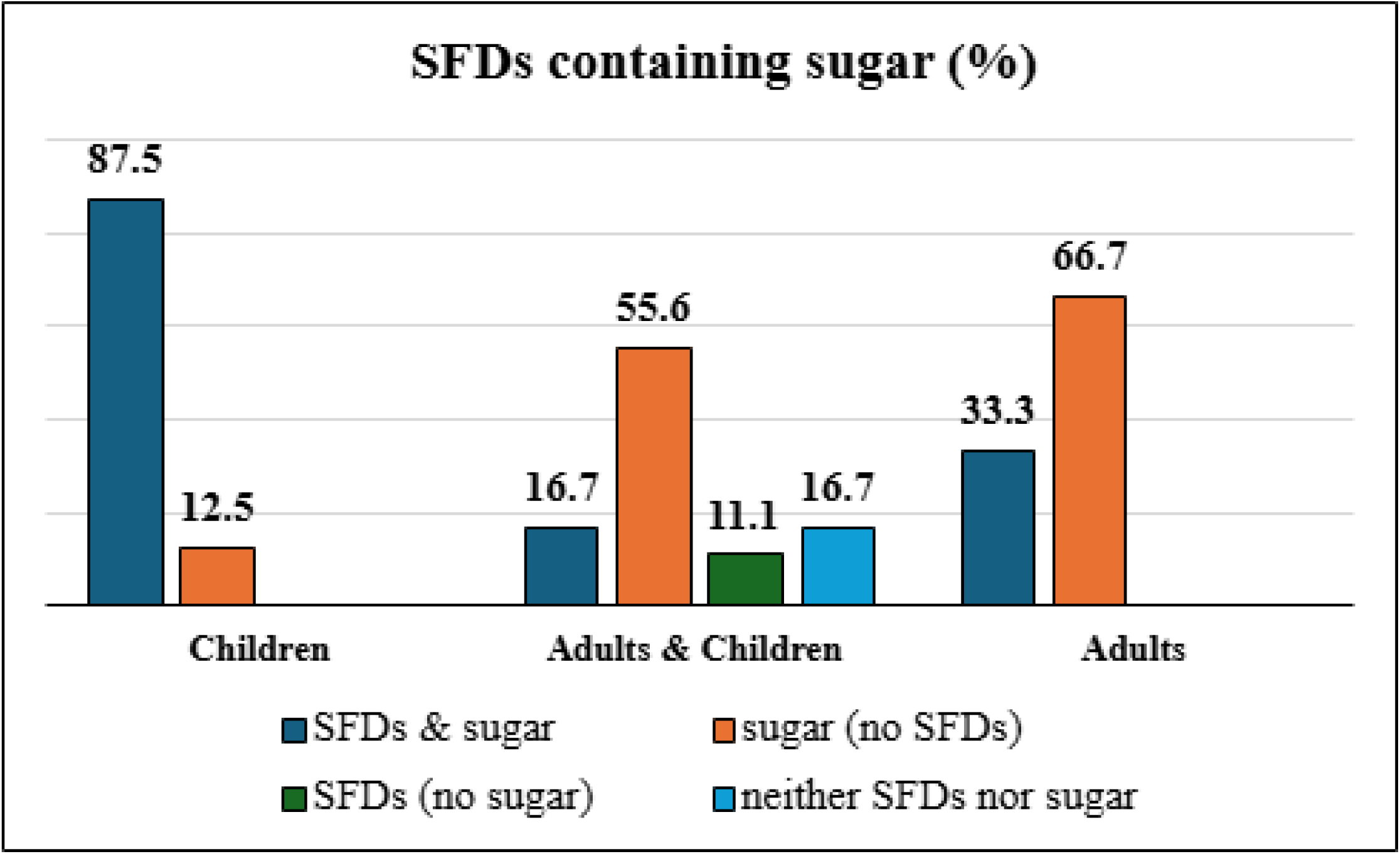
Shows the percentages of each sub-marketing group with specified combinations of SFDs and sugar.

Hydration beverages labeled as “healthy” were less likely to contain SFDs (67.4%). Only 9 (23.6%) of products labeled as healthy for all ages contained SFDs, compared with 38 (76.3%) of all-age products with a label designating health benefits. In contrast, hydration beverages labeled as healthy and marketed for the pediatric population showed a much higher prevalence of SFDs, with 18 of 20 products (90%) containing SFDs.

**Figure 5.**
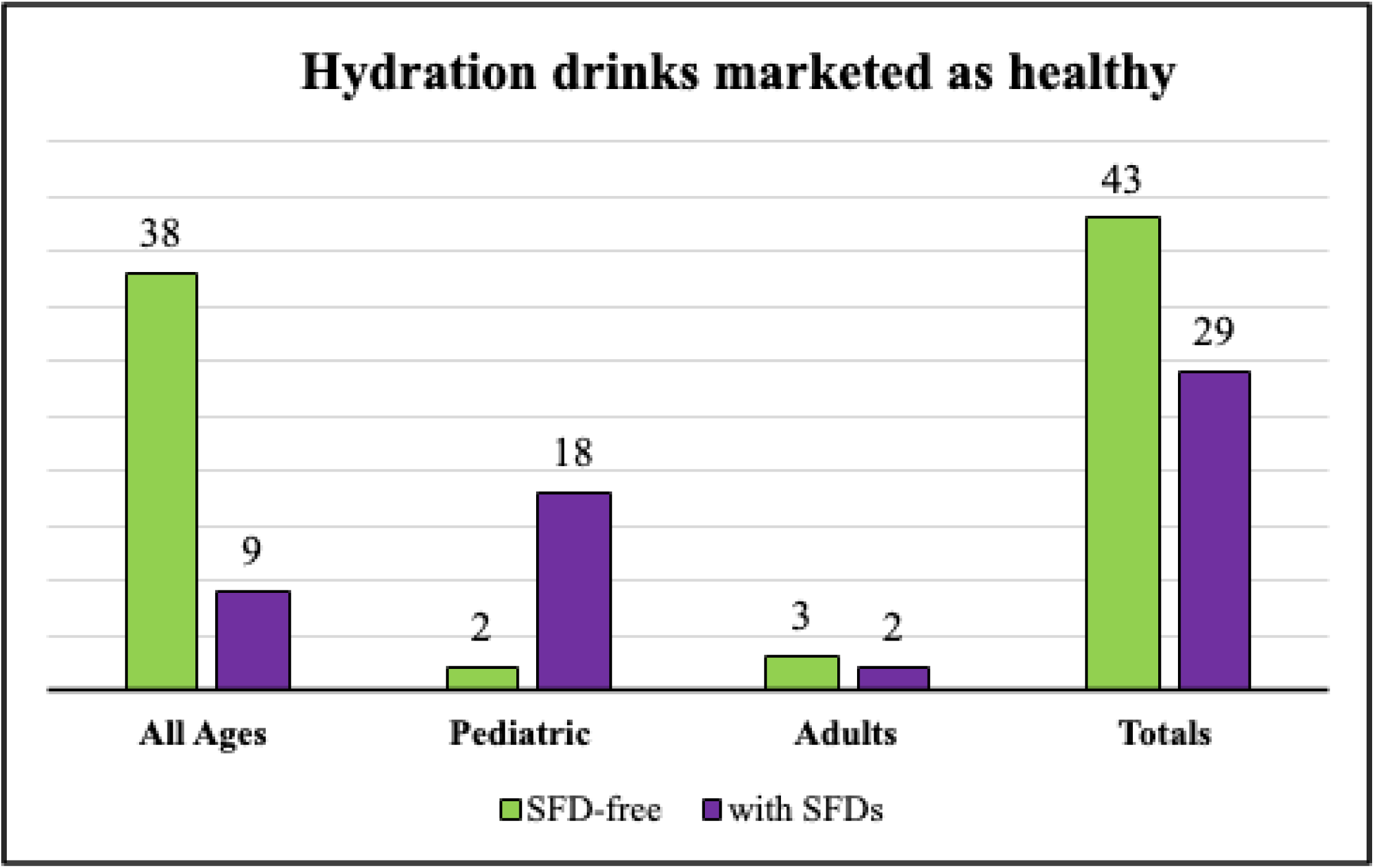
Illustrates totals for each category based on the marketing designation of hydration drinks as healthy.

## Discussion

Overall, this study highlights the widespread use of synthetic food dyes (SFD) in hydration beverages, particularly in products marketed to infants and children and in those containing added sugars. Nearly half of hydration beverages contained at least one synthetic dye, with a higher prevalence in products marketed to pediatric populations. The disproportionate presence of dyes in pediatric-targeted products (87.5%) compared with all-age hydration products (38.4%) suggests a greater likelihood of exposure in pediatric populations. This is concerning given that infants/children have a lower body mass and developing nervous systems, factors that may increase their sensitivity to xenobiotic substances such as synthetic dyes. In the context of prior research linking SFDs to neurobehavioral disturbances, particularly in children, these findings underscore concerns about the widespread inclusion of synthetic dyes in beverages intended for consumption during periods of illness or dehydration.

Two additional findings from this study relate to the presence of added sugars and the influence of “healthy” labeling. Among products containing added sugar, 67.4% also contained synthetic food dyes (SFDs), suggesting that beverages formulated primarily for taste appeal rather than hydration are more likely to include artificial additives. It is understood that sugar can assist with the hydration process; however, the correlation between added sugar and SFDs demonstrates that SFDs are often paired with sugar in drinks. Given that excessive sugar intake has independently been linked to metabolic disorders and behavioral concerns [26], the co-occurrence of sugar and synthetic dyes in hydration beverages may have compounding health implications.

Products marketed as “healthy” were less likely to contain synthetic dyes, indicating that health-focused branding may be associated with efforts to avoid artificial additives. However, a substantial proportion of hydration beverages without health-related labeling (81.0%) did contain synthetic dyes, highlighting a potential gap in consumer awareness. These findings suggest that labeling practices may influence consumer perception of product healthfulness and underscore the importance of clear information for parents and healthcare providers when making hydration recommendations for children [27–30].

## Conclusion

While this study provides insight into the prevalence of synthetic food dyes in hydration beverages, it is limited by its scope, as data were collected from a single retail location in one geographic area. Future studies should include multiple retail locations across different regions to determine whether these patterns are consistent nationwide. Additional research is also needed to examine the potential behavioral and physiological effects of chronic exposure to synthetic dyes from hydration beverages, particularly in vulnerable populations such as children and individuals with preexisting health conditions.

This study contributes to the growing body of research documenting the presence of synthetic food dyes in foods and beverages and may inform future investigations into the long-term health implications of dye consumption and the development of safer alternatives for hydration products.

## Data Availability

All relevant data are within the manuscript and its Supporting Information files.

